# Smoking-Thrombectomy Paradox: A Prospective Single-Center Study on the Impact of Smoking Status on Reperfusion Rates

**DOI:** 10.1101/2025.11.17.25340453

**Authors:** Tzu-Ning Chen, Jyun-Yuan Huang, Te-Yuan Chen, Yu-Ying Wu, Po-Yuan Chen, Jui-Sheng Chen, Cheng-loong Liang, Kang Lu, Hao-Kuang Wang

## Abstract

**Background:** Smoking is a well-established risk factor for acute ischemic stroke, but its influence on endovascular thrombectomy outcomes remains unclear. While previous studies suggested a “smoking paradox” in thrombolysis, evidence regarding EVT is limited.

**Methods:** We conducted a prospective, single-center observational study of 169 patients with acute ischemic stroke who underwent EVT between January 2019 and March 2023, using data from a tertiary hospital thrombectomy database in Kaohsiung, Taiwan. Patients were categorized as smokers (n=52) or non-smokers (n=117). Baseline characteristics were adjusted using inverse probability of treatment weighting. The primary endpoint was complete reperfusion. Secondary outcomes included NIHSS scores at admission and 24 hours post-procedure, modified Rankin Scale at discharge, and complications.

**Results:** Smokers were younger and more often male, with lower rates of atrial fibrillation. After IPTW adjustment, smokers had significantly lower rates of complete reperfusion compared to non-smokers (30% vs. 53%; OR 0.39; 95% CI 0.19–0.79; p=0.01). They also presented with higher NIHSS scores at admission and 24 hours post-thrombectomy (p<0.05), and an increased risk of stroke-associated pneumonia (OR 2.98; p=0.003). No significant difference in discharge mRS was observed.

**Conclusions:** Smoking is associated with reduced reperfusion success, worse neurological status, and higher pneumonia risk after EVT, but not with short-term functional outcome. These findings contradict the notion of a smoking–reperfusion paradox in EVT-treated AIS patients.

## Introduction

Acute ischemic stroke (AIS) is a leading cause of death and long-term disability worldwide. Endovascular thrombectomy (EVT) has become the standard treatment for large vessel occlusion, providing high rates of reperfusion and improved clinical outcomes. Smoking is a well-established risk factor for AIS, yet its impact on reperfusion therapies remains controversial. Previous studies have described the so-called “smoking paradox,” in which smokers appeared to achieve higher recanalization rates and better outcomes following intravenous thrombolysis^1,2^. However, there is little study mentioning the evidence regarding the effect of smoking on EVT outcomes.

The TREAT-AIS^3^ study is a large multicenter, prospective, observational registry conducted in Taiwan, involving 10 medical centers and 9 community hospitals, including our institution. This nationwide collaboration enrolled patients aged 20 years or older who underwent EVT for acute ischemic stroke and collected detailed clinical information such as vascular risk factors, laboratory findings, periprocedural complications, neurological assessments, and functional outcomes. Given the richness of this dataset, it provides a unique opportunity to explore clinically relevant questions. In the present study, we analyzed data from our hospital and focused on the relationship between cigarette smoking and EVT outcomes, aiming to clarify whether smoking status influences reperfusion success and clinical recovery after stroke.

## Methods

### Study Design and Population

This is a prospective, single-centered, observational cohort study conducted at the Center for Stroke Research at Eda hospital. Acute stroke patients who received thrombectomy between January 2019 and March 2023 were enrolled in this study. The inclusion criteria were as follows (1) adult patients (2) thrombectomy performed after acute ischemic stroke. Patients with incomplete data were excluded.

All patients had provided informed consent for medical treatment prior to their inclusion in our hospital’s thrombectomy database, in which all data were anonymized. Because the present study utilized only anonymized data from this database, the requirement for additional informed consent was waived. The study was conducted in accordance with the principles of the Declaration of Helsinki and was approved by the Institutional Review Board of Eda Hospital.

### Clinical Data Collection

An electronic data registration sheet was used for this study. The data sheet included the following variables: age, sex, body mass index (BMI), comorbidities (including hypertension, diabetes mellitus, dyslipidemia, cerebrovascular accident, transient ischemic attack, and atrial fibrillation), smoking history, alcohol consumption history, target vessels, National Institutes of Health Stroke Scale (NIHSS) score at admission and at 24 hours post-thrombectomy, modified Rankin Scale (mRS) score at discharge, and complications.

### Clinical Assessment and Procedures

On admission, each patient was systematically evaluated for cerebrovascular risk factors, including smoking, arterial hypertension, diabetes mellitus, dyslipidemia, atrial fibrillation, chronic kidney disease (CKD), history of transient ischemic attack (TIA), and alcohol consumption. Patients who reported active cigarette use were classified as smokers, whereas those who had quit smoking for more than six months were considered non-smokers.

For patients presenting with stroke-related symptoms, such as headache, hemiplegia, facial drooping, or speech difficulties, diagnostic imaging with brain computed tomography angiography (CTA) and computed tomography reperfusion (CTP), or brain magnetic resonance imaging (MRI), was performed to confirm the diagnosis of acute ischemic stroke. Eligibility for EVT was determined according to the current recommendations of the American Heart Association/American Stroke Association and the Taiwan Stroke Society guidelines.

### Outcomes

The primary endpoint was successful complete reperfusion, defined as a Thrombolysis in Cerebral Infarction (TICI) grade 3. Secondary endpoints included the NIHSS score at admission and 24 hours after thrombectomy, the mRS score at discharge, and the occurrence of complications such as pneumonia, urinary tract infection (UTI), sepsis, acute coronary syndrome (ACS), and seizure. NIHSS was treated as a continuous covariate in the regression analyses. The mRS was dichotomized into favorable (0∼2) and unfavorable (3∼6) outcomes.

### Statistical Analysis

Group comparisons were performed using the chi-square test for categorical variables and the Mann–Whitney U test for continuous variables. To address potential selection bias, we applied inverse probability of treatment weighting (IPTW) to control for differences in baseline characteristics between smokers and non-smokers. Propensity scores were estimated using logistic regression models based on age and sex. Each individual in the smoking group was weighted by 1/propensity score, and those in the non-smoking group were weighted by 1/(1 − propensity score). Covariate balance after weighting was assessed using standardized differences and visualized with Love plots.

Logistic regression and descriptive analyses were conducted to assess the association between smoking status and successful complete reperfusion. NIHSS scores at hospitalization and 24 hours post-procedure were treated as continuous variables and analyzed using univariate and multivariate linear regression. The mRS at discharge was dichotomized and evaluated with logistic regression. Complications were compared using the chi-square test. Two-tailed p values were reported, with statistical significance defined as p ≤ 0.05. All statistical analyses were performed with SPSS Statistics for Windows, version 31.0 (IBM Corp., Armonk, NY, USA).

## Results

### Patient Characteristics

A flowchart of participant selection is presented in (Figure 1). A total of 184 patients who underwent mechanical thrombectomy for acute ischemic stroke were initially enrolled. After excluding patients with incomplete data, 169 patients were included in the final analysis. Among these, 52 (30.8%) were classified as the smoking group, and 117 (69.2%) as the non-smoking group. Baseline characteristics of the two groups are summarized in (Table 1).

**Table 1.** Baseline Characteristics of Patients.

| Variable | Before IPTW Adjustment |  |  | After IPTW Adjustment |  |  |
| --- | --- | --- | --- | --- | --- | --- |
|  | Smoker<br>(n=52) | Non-smoker<br>(n=117) | p-value* | Smoker | Non-smoker | p-value* |
| Age (years) | 59.46 ± 1.9 | 73.96 ± 2.03 | <0.001 | 68.29 ± 2.07 | 68.65 ± 1.3 | 0.707 |
| BMI (kg/m<sup>2</sup>) | 24.92 ± 0.54 | 24.47 ± 3.37 | 0.334 | 25.15 ± 0.49 | 25.41 ± 0.43 | 0.745 |
| Sex male | 44 (84.6%) | 42 (35.9%) | <0.001 | 60% | 53% | 0.409 |
| Hypertension | 28 (53.8%) | 91 (77.8%) | 0.002 | 72% | 69% | 0.694 |
| Diabetes mellitus | 15 (28.8%) | 45 (38.5%) | 0.228 | 28% | 33% | 0.598 |
| Dyslipidemia | 15 (28.8%) | 38 (32.5%) | 0.639 | 24% | 30% | 0.436 |
| CVA | 6 (11.5%) | 24 (20.5%) | 0.286 | 13% | 18% | 0.415 |
| TIA | 0 (0%) | 2 (1.7%) | 0.638 | 0% | 0.8% | 0.68 |
| Atrial fibrillation | 13 (25.0%) | 56 (47.9%) | 0.005 | 22% | 41% | 0.021 |
| Alcohol use | 26 (50.0%) | 5 (4.3%) | <0.001 | 43% | 6% | <0.001 |
| Target Vessel<br>(Anterior Circulation) | 47 (90%) | 101 (86%) | 0.46 | 94% | 86% | 0.178 |

Data before weighting are presented as number (%) or mean ± SD. Post-IPTW adjustment cohort data are presented as mean ± SD or %. The total numbers of participants in the IPTW columns were omitted as the patient numbers were altered because of weighting.
\* Determined with the Pearson's chi-square test and Mann-Whitney U test.

**Figure 1.**
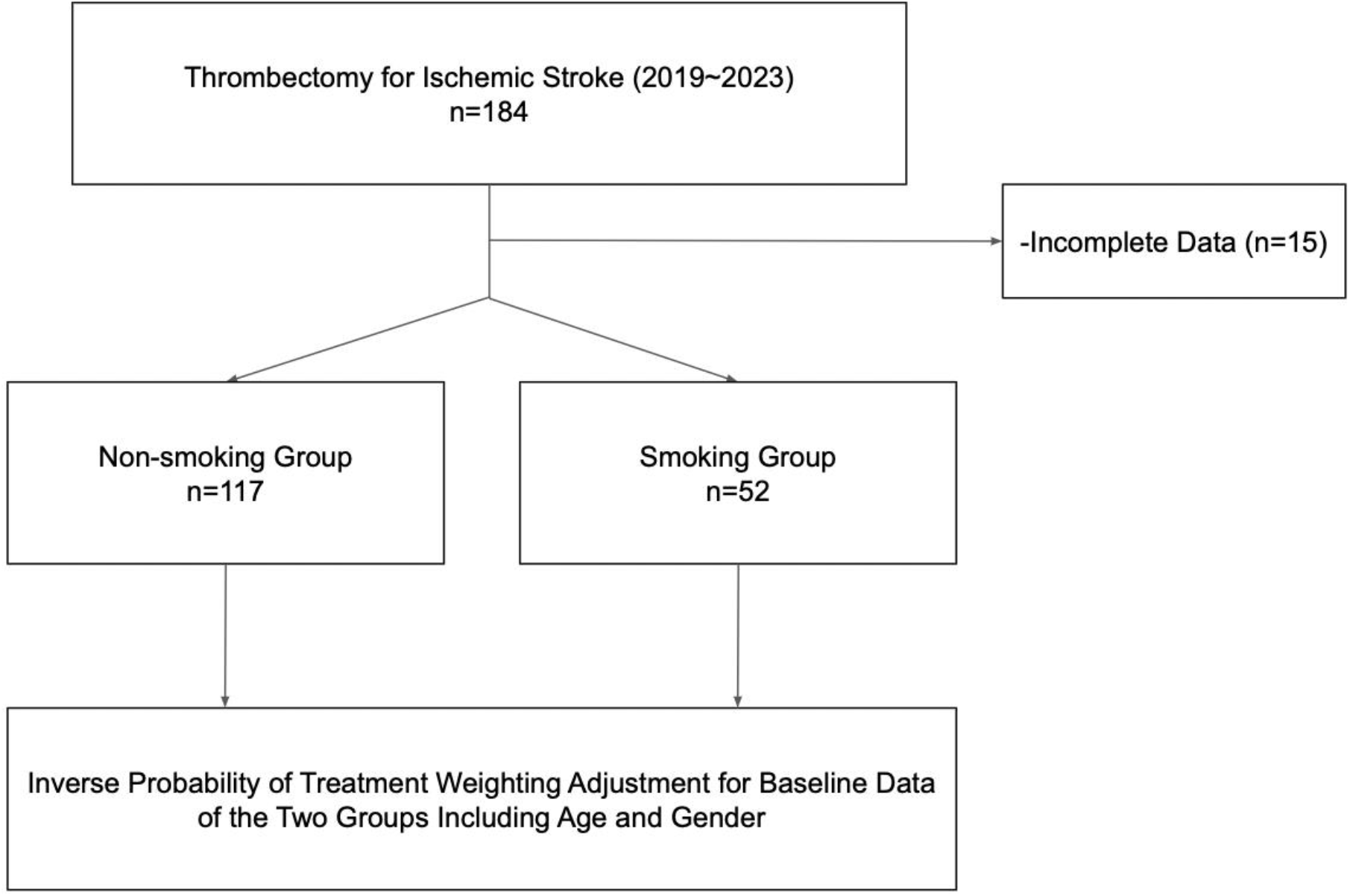
Study Flowchart: A total of 184 patients who underwent mechanical thrombectomy for acute ischemic stroke were initially enrolled. After excluding patients with incomplete data, 169 patients were included in the final analysis. Among these, 52 (30.8%) were classified as the smoking group, and 117 (69.2%) as the non-smoking group.

Compared with the non-smoking group, the smoking group was significantly younger (mean age, 59.46 vs. 73.96 years; p < 0.001), had a higher proportion of male patients (84.6% vs. 35.9%; p < 0.001), more alcohol use (50.0% vs. 4.3%; p < 0.001), and a lower prevalence of hypertension (53.8% vs. 77.8%; p = 0.002). Atrial fibrillation was less common in the smoking group (25.0% vs. 47.9%; p = 0.005). There were no significant differences between groups in body mass index (BMI), diabetes mellitus, dyslipidemia, history of cerebrovascular accident (CVA), or transient ischemic attack (TIA). Additionally, there was no significant difference in the distribution of target vessels between the anterior and posterior circulations.

To address baseline differences between the two groups, inverse probability of treatment weighting (IPTW) was applied using age and sex, both considered clinically relevant covariates. After IPTW adjustment, the standardized mean difference (SMD) for age was 0.025, and the SMD for sex was 0.14—slightly above the conventional threshold of 0.1 but substantially improved compared to the pre-IPTW value (Figure 2). All subsequent analyses were performed using IPTW-adjusted data.

**Figure 2.**
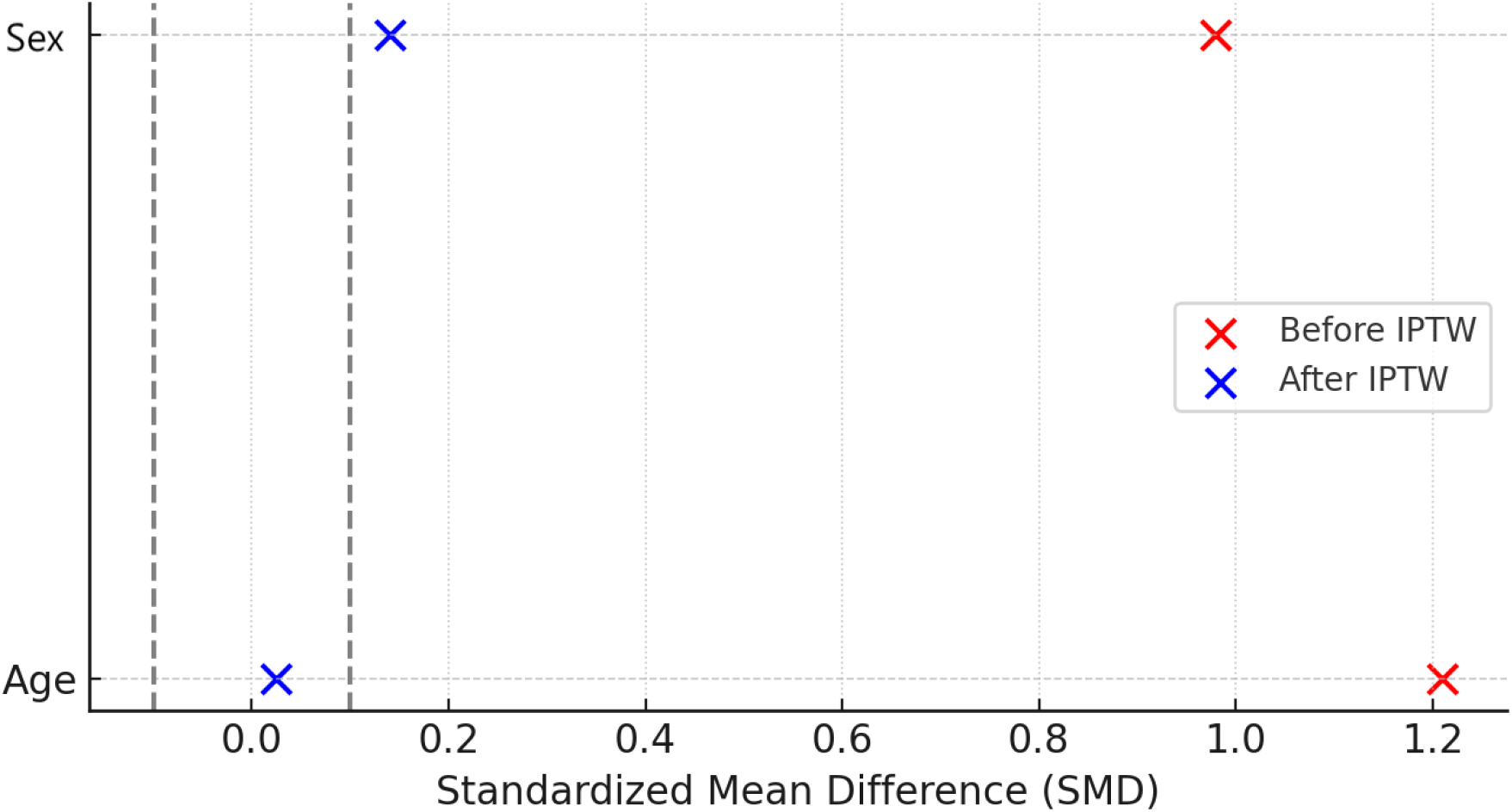
Love plot of Covariate balance: After IPTW adjustment, the SMD for age was 0.025, and the SMD for sex was 0.14—slightly above the conventional threshold of 0.1 but substantially improved compared to the pre-IPTW value.

### Primary Endpoint

Patients who achieved grade 3 on the TICI scale were considered to have reached the primary endpoint. Compared with the non-smoking group, patients in the smoking group had a significantly lower rate of complete reperfusion (30% vs. 53%; OR, 0.39; 95% CI, 0.19–0.79; p = 0.01) (Table 2, Figure 3).

**Table 2.** Success Rate of Complete Reperfusion based on smoking status.

|  | Success Rate |  | Odds ratio (95%CI) | p value |
| --- | --- | --- | --- | --- |
|  | Non-smokers | Smokers |  |  |
| Complete Perfusion | 0.53 | 0.3 | 0.39 (0.19–0.79) | 0.01 |

**Table 3.** Response of Clinical Outcomes to thrombectomy based on smoking status:

|  | Univariate Analysis |  | Multivariate Analysis† |  |
| --- | --- | --- | --- | --- |
| | OR/ $\beta$ coefficient<br>(95% CI) | p value | $\beta$ coefficient<br>(95% CI) | p value |
| NIHSS_Hospitalization | -2.95 (-5.23,-0.67)* | 0.012 | -2.98 (-5.27~-0.68) | 0.011 |
| NIHSS_24hr | -4.77 (-8.25, -1.29)* | 0.007 | -4.93 (-8.37~-1.49) | 0.005 |
| mRS_Discharge | 0.34 (0.52~3.81)† | 0.5 | - | - |

NIHSS was treated as continuous variables and analyzed with univariate and multivariate regression. The mRS at discharge was dichotomized into favorable (0–2) and unfavorable (3–6) outcomes and analyzed with logistic regression model.
\* $\beta$ coefficient; † odds ratio

**Table 4.** Complications.

|  | Incidence |  | RR (95%CI) | Odds ratio (95%CI) | p value |
| --- | --- | --- | --- | --- | --- |
|  | Non-Smokers | Smokers |  |  |  |
| All | 0.57 | 0.66 | 1.11 (0.92~1.34) | 1.48 (0.73~2.99) | 0.272 |
| Pneumonia | 0.29 | 0.54 | 1.36 (1.09~1.74) | 2.89 (1.43~5.83) | 0.003 |
| Sepsis | 0.06 | 0.02 | 0.82 (0.62~1.08) | 0.36 (0.43~2.99) | 0.324 |
| UTI | 0.22 | 0.35 | 1.23 (0.98~1.98) | 1.93 (0.91~4.07) | 0.082 |
| ACS | 0.02 | 0.01 | 0.72 (0.65~0.79) | 0.36 (0.01~10.93) | 0.279 |
| Seizure | 0.05 | 0.04 | 0.96 (0.64~1.45) | 0.86 (0.17~4.44) | 0.861 |

**Figure 3.**
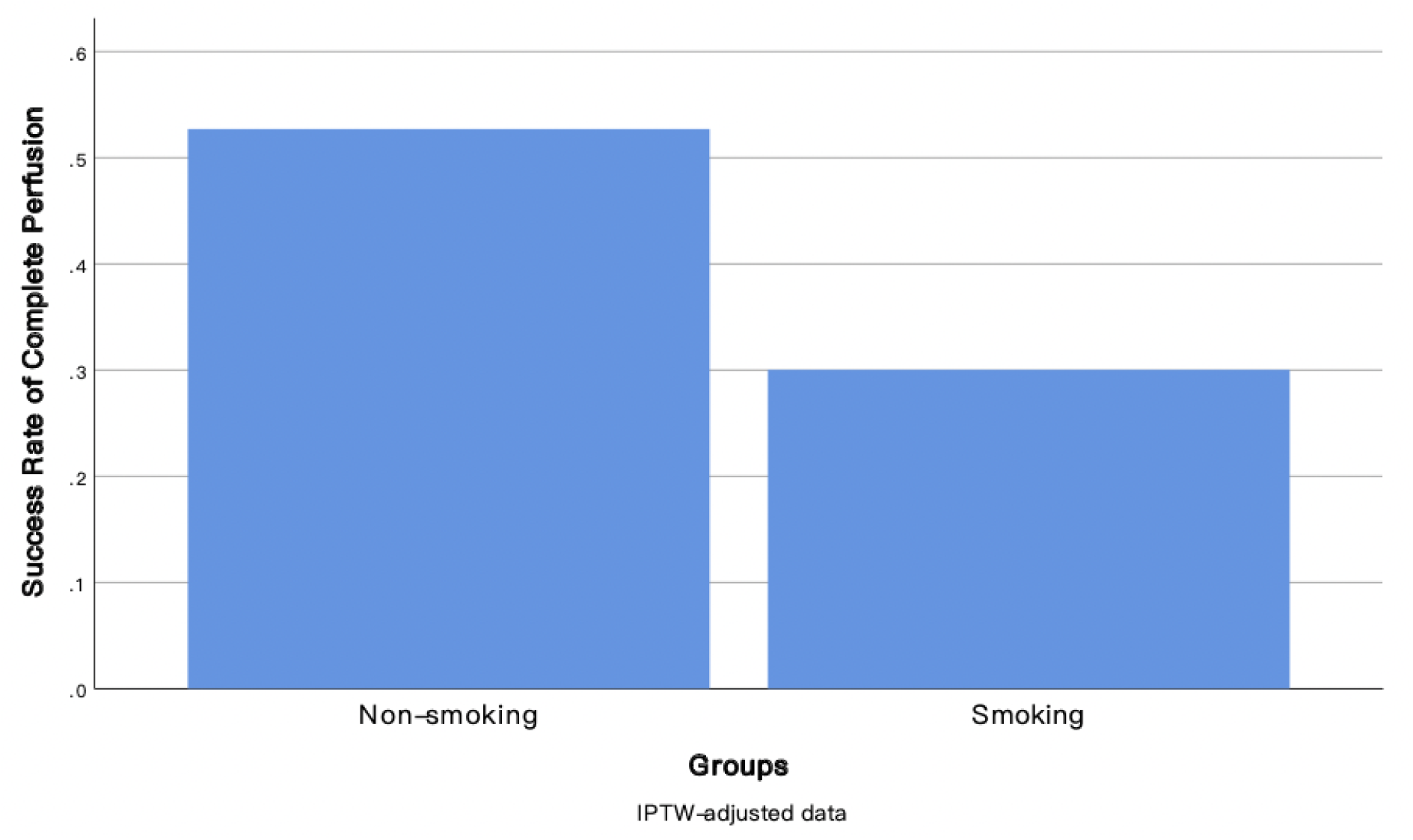
Bar Chart of Success Rate of Complete Reperfusion between Smoking and Non-smoking Groups

**Figure 4.**
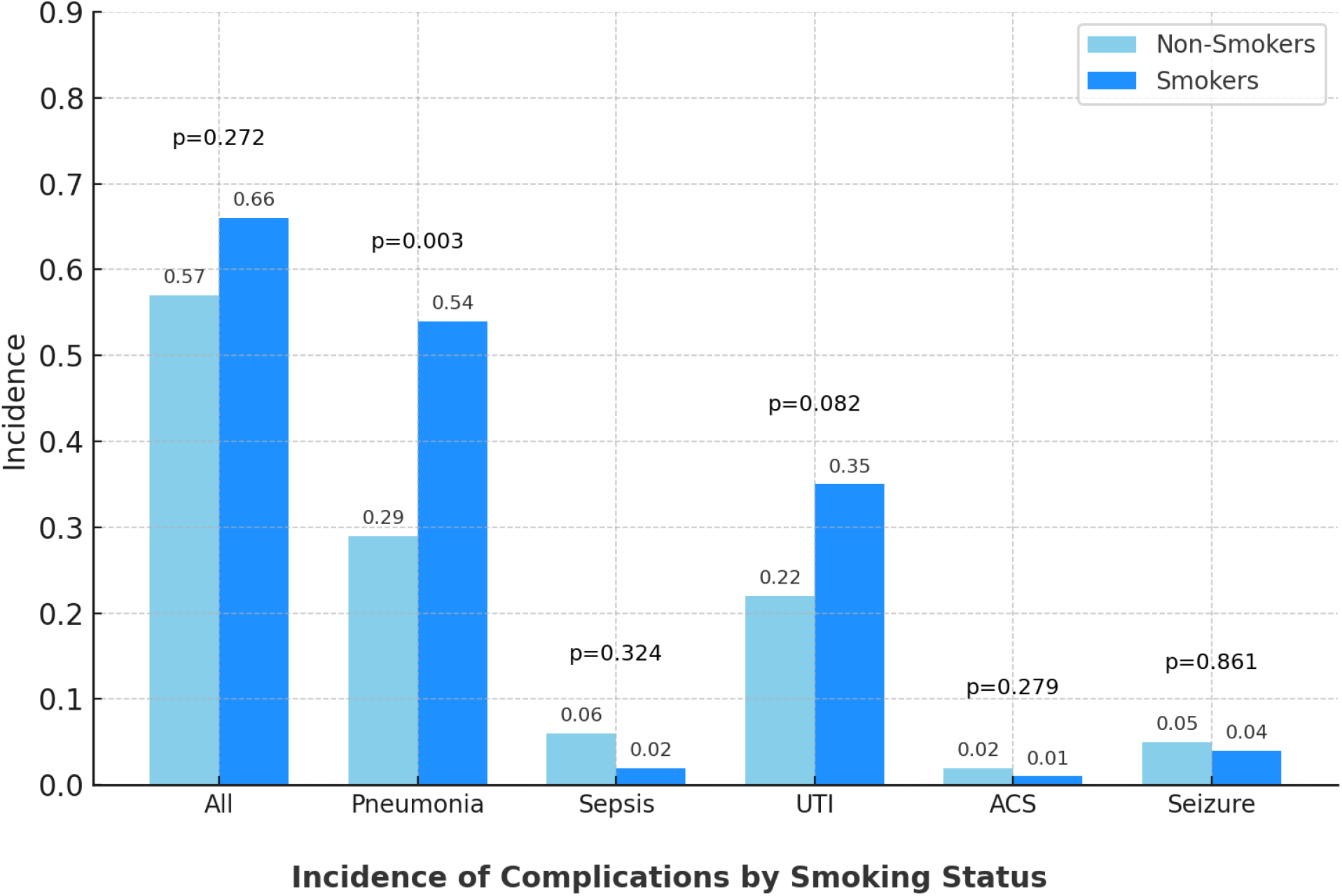
Complications.

### Secondary Endpoint

For secondary endpoints, the association between smoking and clinical outcomes was further examined. Smokers had significantly higher NIHSS scores at admission (mean difference 2.98; 95% CI 0.67–5.23; p= 0.012) and at 24 hours after thrombectomy (mean difference 4.77; 95% CI 1.29–8.25; p= 0.007) compared with non-smokers. Multivariate regression confirmed these findings, showing consistently higher NIHSS scores in smokers at both hospitalization (p= 0.011) and 24 hours post-procedure (p= 0.005). The mRS at discharge was analyzed as an ordinal variable and further dichotomized into favorable (0–2) and unfavorable (3–6) outcomes. Logistic regression analysis demonstrated no significant association between two groups.

Complications were compared between groups using the chi-square test. Smokers had a significantly higher incidence of pneumonia compared with non-smokers (OR 2.98, 95% CI 1.43∼5.85, p=0.003), whereas other complications, including sepsis, urinary tract infection, acute coronary syndrome, and seizure, were not significantly associated with smoking status.

## Discussion

In this prospective, single-center study of 169 patients with acute ischemic stroke, the primary endpoint of complete reperfusion was achieved in 53% of non-smokers compared with 30% of smokers. Smokers had significantly lower odds of achieving complete reperfusion (OR, 0.39; 95% CI, 0.19–0.79; p = 0.01). Clinical outcomes were further evaluated using two scales. Compared with non-smokers, smokers had significantly higher NIHSS scores at admission (β = –2.95; 95% CI, –5.23 to –0.67; p = 0.012) and at 24 hours after thrombectomy (β = –4.77; 95% CI, –8.25 to –1.29; p = 0.007). No significant difference in functional outcome was observed when assessed by the mRS at discharge (OR, 0.34; 95% CI, 0.52∼3.81; p = 0.5)

To our knowledge, this is the first study to investigate the association between smoking and outcomes of EVT in patients with acute ischemic stroke. The so-called smoking–reperfusion paradox has been a matter of debate for a long time. Kufner et al.^2^ reported that smoking was independently associated with enhanced tPA efficacy and higher rates of recanalization and reperfusion. Kurmann et al.^1^ found smoking to be significantly associated with higher recanalization rates in patients with M1 occlusion treated with intravenous thrombolysis. In contrast, our cohort demonstrated that smoking was significantly associated with lower rates of recanalization following EVT. This discrepancy may be explained by differences in thrombus composition and vascular pathology^4^. Compared with non-smokers, thrombi from smokers are richer in fibrinogen and platelets and more responsive to rt-PA, which binds to fibrin and facilitates clot lysis^4^. Chronic smoking also promotes vascular remodeling, endothelial dysfunction, and increased arterial stiffness^4,5^. Moreover, smoking has been reported to reduce cerebral blood flow and increase blood viscosity^6^. Collectively, these factors may create a less favorable endovascular environment, complicating EVT by hindering device navigation and increasing the difficulty of thrombus retrieval.

Xu et al^7^. reported that smoking was not associated with favorable 90-day outcomes following intravenous thrombolysis. Similarly, Zhang et al.^8^ identified smoking as a risk factor for poor 3-month outcomes in patients with severe AIS, and Matsuo et al.^9^ found current smoking to be associated with an increased risk of unfavorable functional outcomes at 3 months after acute ischemic stroke. In our study, after adjusting for confounders, non-smokers demonstrated significantly better neurological function, as reflected by lower NIHSS scores at admission (mean difference, 2.98 points; 95% CI, 0.67–5.23; p = 0.012) and at 24 hours after thrombectomy (mean difference, 4.77 points; 95% CI, 1.29–8.25; p = 0.007). These findings suggest that smoking has a detrimental effect on baseline and early post-thrombectomy neurological status. However, no significant difference was observed between groups with respect to functional outcomes at discharge as measured by the mRS. This discrepancy may be explained by the limited 7-point range of the mRS (0–6) compared with the broader 43-point range of the NIHSS (0–42). Notably, a previous study demonstrated that 24-hour NIHSS is a reliable surrogate for 90-day mRS in patients with AIS undergoing EVT^10^.

As expected, smoking was associated with younger age, male sex, lower prevalence of atrial fibrillation, and higher alcohol consumption. After IPTW adjustment, non-smokers continued to show a significantly higher rate of atrial fibrillation, which may partly explain their increased risk of AIS. In our cohort, the mean age at stroke onset was 59.5 years in smokers and 73.9 years in non-smokers. These findings are consistent with a previous study reporting mean onset ages of 60.2 years for current smokers and 71.6 years for non-smokers^11^. The age difference between the two groups in our study was approximately 14 years, indicating that smokers experience stroke, along with its associated increasing neurological deficits and disability, more than a decade earlier than non-smokers. The overall smoking rate in this study is around 30%, relatively higher than the adult smoking rate in Taiwan, which is around 14% in 2022. This may be attributable to the hospital’s location in an industrial area.

Stroke-associated pneumonia (SAP) is one of the most frequent and serious complications following stroke. In our study, the incidence of SAP was 36%, which is higher than the 13–23% reported in previous studies^12-14^. This discrepancy may in part be attributable to the higher prevalence of smoking in our cohort. Our study also demonstrated that smokers had a higher risk of developing SAP. SAP is primarily related to aspiration, as patients with AIS often experience neurological deficits such as impaired swallowing and a weakened cough reflex. In addition, the cholinergic pathway has been implicated in the pathophysiology of SAP. Experimental studies demonstrated rapidly stimulated parasympathetic activity after stroke in mice, leading to impaired alveolar immune responses and increased susceptibility to pneumonia^15^. Patients who develop SAP typically have worse prognoses, including higher mortality and poorer long-term outcomes^12,13^. Prophylactic antibiotic therapy is not recommended, as it has been shown to have no impact on SAP incidence, underscoring the importance of prevention strategies focused on attenuated cough function and dysphagia after stroke^14^.

This study has some limitations. First, the relatively small sample size may introduce selection bias and limit statistical power, increasing the risk of type II errors. Secondly, although IPTW was applied to adjust for baseline confounders including age and sex, the SMD for sex after weighting remained 0.14, indicating residual imbalance. Due to limited sample size, we could only include the age and sex in the propensity score model. Consequently, we cannot entirely rule out the potential influence of other unmeasured confounding variables. As a result, the results should be interpreted with caution. There was a lack of quantitative data including how many cigarettes in one day and how long the smoking history persisted. Additional analyses of trials that include larger size of data with more detailed information of smoking behavior would be excellent to explore this topic further.

## Conclusions

Our study demonstrated that smoking is an unfavorable predictor of complete reperfusion after thrombectomy in patients with acute ischemic stroke. Smoking is also associated with worse neurological status and a higher risk of stroke-associated pneumonia, although it does not significantly affect functional outcomes at discharge. In conclusion, our findings do not support the existence of a smoking–reperfusion paradox in patients treated with thrombectomy.

## Data Availability

all of data belong to e da hospital

## Funding

This study did not receive any funding or other financial support.

## Disclosure

The authors declare that they have no competing interests.

## Notes

### Competing Interest Statement

The authors have declared no competing interest.

### Clinical Trial

No

### Funding Statement

e da hospital support

### Author Declarations

The study was conducted in accordance with the principles of the Declaration of Helsinki and was approved by the Institutional Review Board of Eda Hospital. EMRP12106N

